# Investigating the comparability of wearable accelerometer methods in the association between physical activity and cardiovascular disease: a cohort study using UK Biobank

**DOI:** 10.64898/2026.01.07.26343633

**Authors:** Yacine Lapointe, Aayush Kapur, Abhinav Sharma, Janna Panter, Daniel Fuller, Hiroshi Mamiya

## Abstract

**Objective:** The selection of accelerometer processing methods may influence the shape of the dose-response association between wearable-measured physical activity and health outcomes. We aimed to compare the association of stroke and myocardial infarction with Moderate-Vigorous Physical Activity (MVPA) assessed by three accelerometer-generated metrics: Low-pass Filtered Euclidean Norm Minus One (LFENMO), machine-learning, and activity counts.

**Methods:** We computed MVPA durations in the UK Biobank accelerometer sub-cohort recruited between 2013 and 2015 in the UK. The outcomes, incident stroke and myocardial infarction, were followed up until December 2022. We used Cox regression and a restricted cubic spline to estimate the dose-response association for each of the three MVPA metrics.

**Results:** There were 90,237 cardiovascular disease-free participants at baseline. We observed 1,298 incident strokes and 2,031 myocardial infarctions. For stroke, a linear decrease in hazard ratio was observed with machine-learning, but not with LFENMO and activity counts. For myocardial infarction, machine-learning and LFENMO showed a curvilinear decrease in hazard ratios, whereas activity counts showed a linear decrease.

**Conclusions:** The dose-response associations between MVPA and cardiovascular disease varied markedly across the three accelerometer-derived MVPA metrics. Research using a single accelerometer metric may caution about the interpretation of the association.

## 1. Background

Insufficient Physical Activity is the major risk factor for Cardiovascular Disease (CVD)(1–3). The national and international physical activity guidelines recommend greater than 150 minutes/week of Moderate to Vigorous Physical Activity (MVPA)(4,5). MVPA is the intensity of activity enough to elevate the heart rate, including brisk walking (moderate) and running (vigorous) (6). While meeting this recommendation can reduce CVD-related incidence, nearly 29% of males and 34% of females failed to meet this recommendation globally (7).

A meaningful reduction in the risk of CVD may occur well below the 150 minutes/week threshold of MVPA, with the benefit starting as low as 60 minutes/week (8). Accurate characterization of the dose-response association between CVD and physical activity is critical to assess the population burden of insufficient activity, model the reduction in CVD by shifting population levels of physical activity, and provide refined physical activity recommendations.

Existing meta-analyses agree on the incomplete understanding of the dose-response association due to the inaccuracy of physical activity measures by self-report (9,10). Recent studies using accelerometer-derived activity measures suggest either a curvilinear or linear reduction of CVD risk as the duration of MVPA increases, with varying slopes and plateaus of risk reduction (11–13).

There are numerous approaches to convert device-generated accelerometer data into activity intensity and duration to estimate MVPA(19–21). Researchers commonly compute continuous values of summary metrics within a discrete time window called an epoch from accelerometer data (14,15). These summary metrics can be computed from raw tri-axial (x/y/z-axes) accelerometer signals, including Monitor-Independent Movement Summary and Euclidean Norm Minus One (16). Some metrics are manufacturer-developed measures, such as activity counts derived from triaxial or single-axis (typically y-axis) accelerometer signals (17). These summary values are often discretized into relevant intensity categories, such as MVPA, light physical activity, and sedentary activity, at the epoch level, before aggregating to obtain the total time spent in each intensity category. Such categorization is based on pre-defined thresholds or ‘cut-points’ derived from calibration studies utilizing ground-truth data specific to age and device wear locations (e.g., hip and wrist) (14,18). There are also methods that do not rely on pre-determined cut-points, including machine-learning. Machine-learning can estimate activity intensity at each epoch using numerous predictive feature variables computed from raw accelerometer data or summary metrics (e.g., activity counts) (17,19).

The selection of accelerometer methods influences study results. Comparison studies showed discrepancies in the prevalence of meeting the PA guidelines within the same populations due to the varying accuracy of these methods to quantify MVPA (14,20). There is limited knowledge about how decisions in selecting accelerometer methods influence dose-response associations between accelerometer-derived PA and health outcomes. Such knowledge will help explain the heterogeneity in dose-response associations and promote the standardization and harmonization of derived physical activity (20).

The primary objective of this study was to compare dose-response associations between CVD (incident stroke and myocardial infarction) across three accelerometer metrics of MVPA from the UK Biobank accelerometer cohort. These are Low-pass Filter Euclidean Norm Minus One (LFENMO), machine-learning, and activity count-based metrics, with the first and third metrics using previously reported intensity cut-points for MVPA. As a secondary objective, we assessed agreement between the three metrics and ground-truth MVPA.

## 2. Methods and data

### 2.1 Study design and population

This was a longitudinal (time-to-event) study. UK Biobank is a prospective cohort that has been collecting biomedical data from over 500,000 participants aged 40-69 since the baseline assessment in 2006. Our study population was the UK Biobank accelerometer sub-cohort, consisting of 103,686 participants who consented to wear wearable devices for seven consecutive days between 2013 and 2014 (21). Individuals meeting the definition of insufficient wear time (less than 72hours) were excluded (21). We also excluded individuals who had a stroke or myocardial infarction before accelerometer data collection. We also removed those missing baseline covariates, e.g., diets, income, education, and smoking.

### 2.2 Accelerometer Measures

In UK Biobank, wrist-worn Axivity Ax3 (Axivity, United Kingdom) generated raw tri-axial accelerometer data for seven consecutive days. The data were previously processed by calibrating signals to local gravity, removing infeasible accelerometer levels, imputing non-wear time, computing the vector magnitude (i.e., Euclidean Norm) from the triaxial data, and applying a Butterworth low-pass filter to remove noise (21). The resulting five-second (epoch) summary of vector-magnitude acceleration, minus one gravitational unit, from these low-pass-filtered accelerometer signals is provided by UK Biobank as LFENMO (16,21). To compute the time spent on MVPA for each participant in minutes/week, we applied a commonly used and validated cut-point of 100mg to classify MVPA for ENMO (18).

For machine-learning, we used a previously reported machine-learning model using UK Biobank, which combines a random forest and a hidden Markov model (22). The model was previously trained on predictive features from raw accelerometer data at 30-second epochs aligned with ground truth data from 151 participants’ 24-hour activity data called Capture-24 (23). Previous studies already used this machine-learning model to estimate the association of physical activity with CVD using UK Biobank, albeit focusing on the compositional exposure (24-hour activity) or combination with bouts, rather than MVPA and/or with a shorter follow-up time ending in 2021 than our study, extending to the end of 2022 (11,22,23).

For activity counts, we applied an open-source reverse engineering method to reproduce previously manufacturer-proprietary activity counts at 30-second epoch resolution from raw accelerometer data after the pre-processing steps described above (24,25). We then used cut-points for wrist-worn devices validated against free-living adult participants (15). These are <2,860 CPM (Counts Per Minute) for sedentary, 2,860-3,940 CPM for LPA, and ≥ 3,941 CPM for MVPA, multiplied by 0.5 seconds for our 30-second epoch. We note that the algorithm to exactly reproduce activity counts from raw accelerometer data has been available since 2022, rather than the approach that generates approximate counts used in this study (26).

### 2.3 Outcome measures

The two outcomes are the first episode of stroke and myocardial infarction (both fatal and non-fatal), which were ascertained by the UK Biobank’s adjudicated algorithms integrating hospital episodes, death certificates, and self-reports as identified by Field IDs 42009 and 42001, respectively (UK Biobank Resource 460: Algorithmically defined outcomes) (27). Follow-up of participants began after the collection of accelerometer data (between 2013-2014, depending on participants) and continued until death, diagnosis of first stroke or myocardial infarction, or the end of the last available date in the hospital and death records on November 30, 2022.

### 2.4 Statistical analysis

First, we compared participant characteristics by outcome status. We then assessed the agreement between the three MVPA metrics and ground-truth MVPA duration using Bland- Altman plots and Spearman’s rank correlation coefficient (given the non-normal distribution of MVPA) in 151 participants from the Capture-24 validation study. We used the *Accelerometer* Python library to generate a prediction for the validation sample using the pre-trained machine-learning method for MVPA prediction as above, without modification (22,28). . For dose-response analysis, we used Cox regression to model the time to stroke and myocardial infarction separately, with time measured in months since the accelerometer assessment. To account for non-linear associations, we used a restricted cubic spline function with its knots placed at the Fifth, 50^th^, and 95^th^ percentiles of the data (29). We evaluated the number of knots and the linearity (use of the spline or not) based on Akaike’s Information Criterion. We used Schoenfeld residuals to evaluate the proportional hazards assumption.

We added potential confounder variables, including age on a continuous scale and sex, education, and income as categorical variables. We also added self-reported consumption of tobacco smoking, alcohol consumption, red meat, fruits, vegetables, and oily and non-oily fish, all on a categorical scale (11,22). We also controlled for ethnic background (UK Biobank collects ethnicity rather than race), dichotomized into visible minorities (Asian or Asian British, Black or Black British, and other non-White groups) vs. White, given the limited proportion of each non-White group. The categorization of these variables was already defined by UK Biobank (Table 1).

**Table 1.**
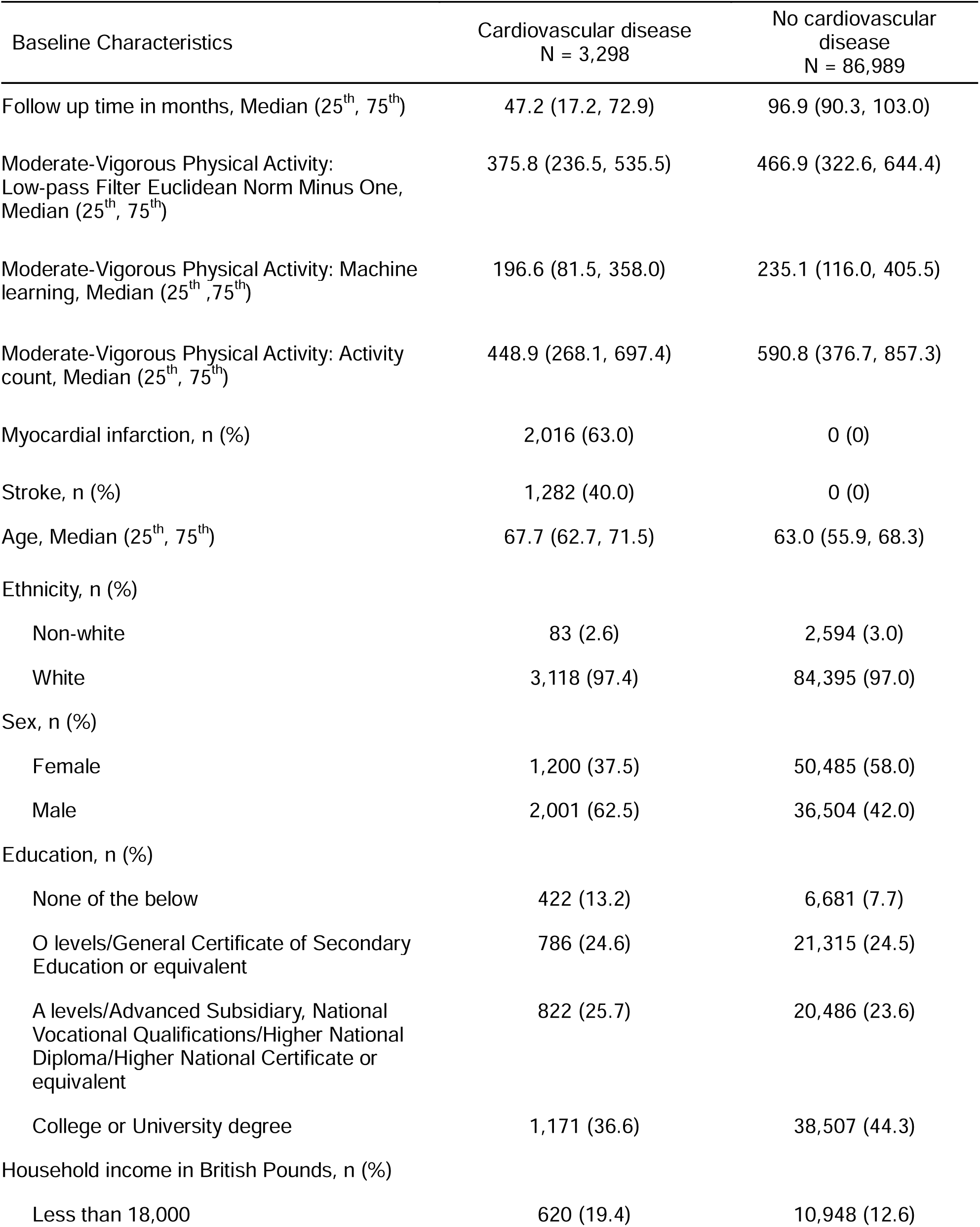
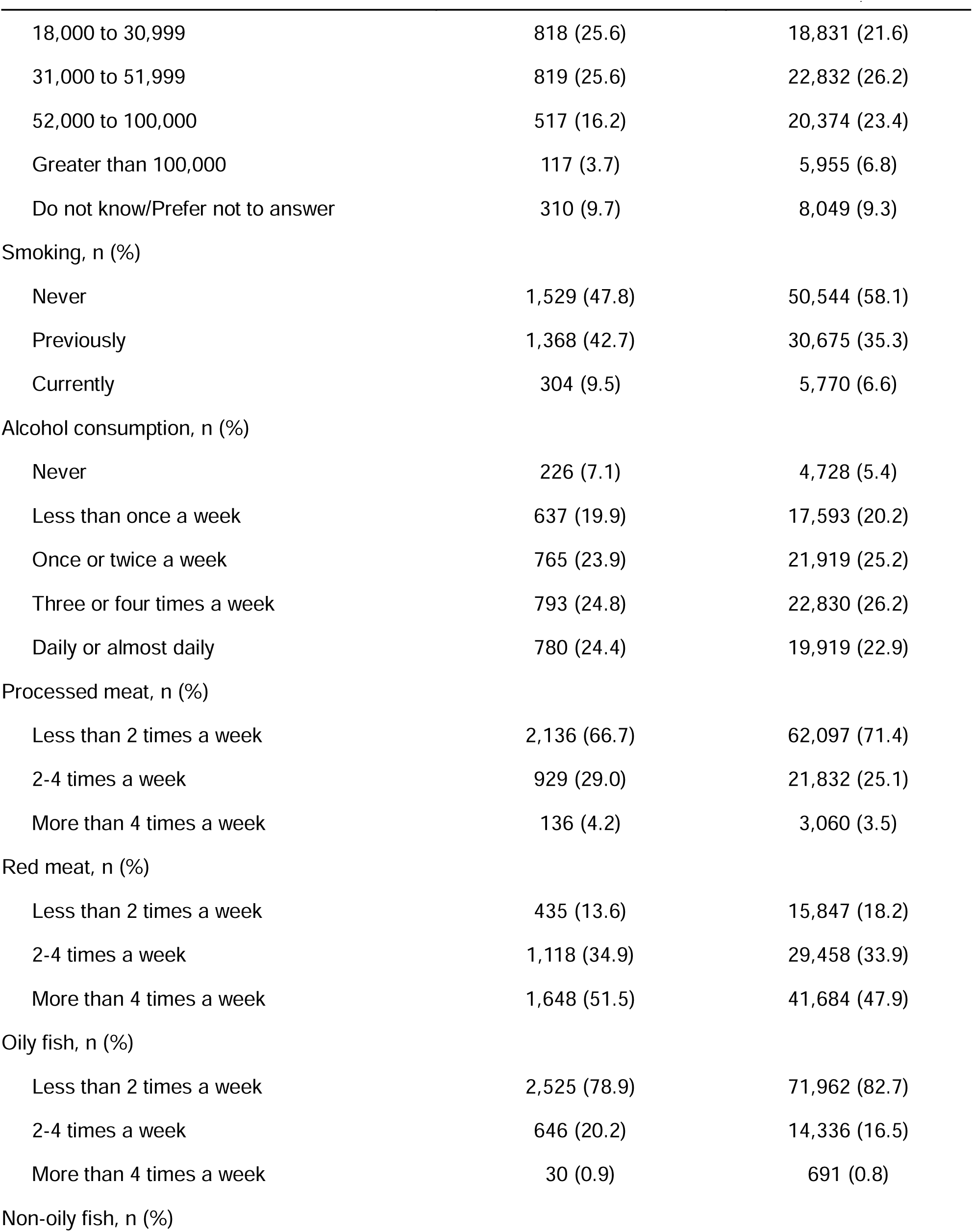
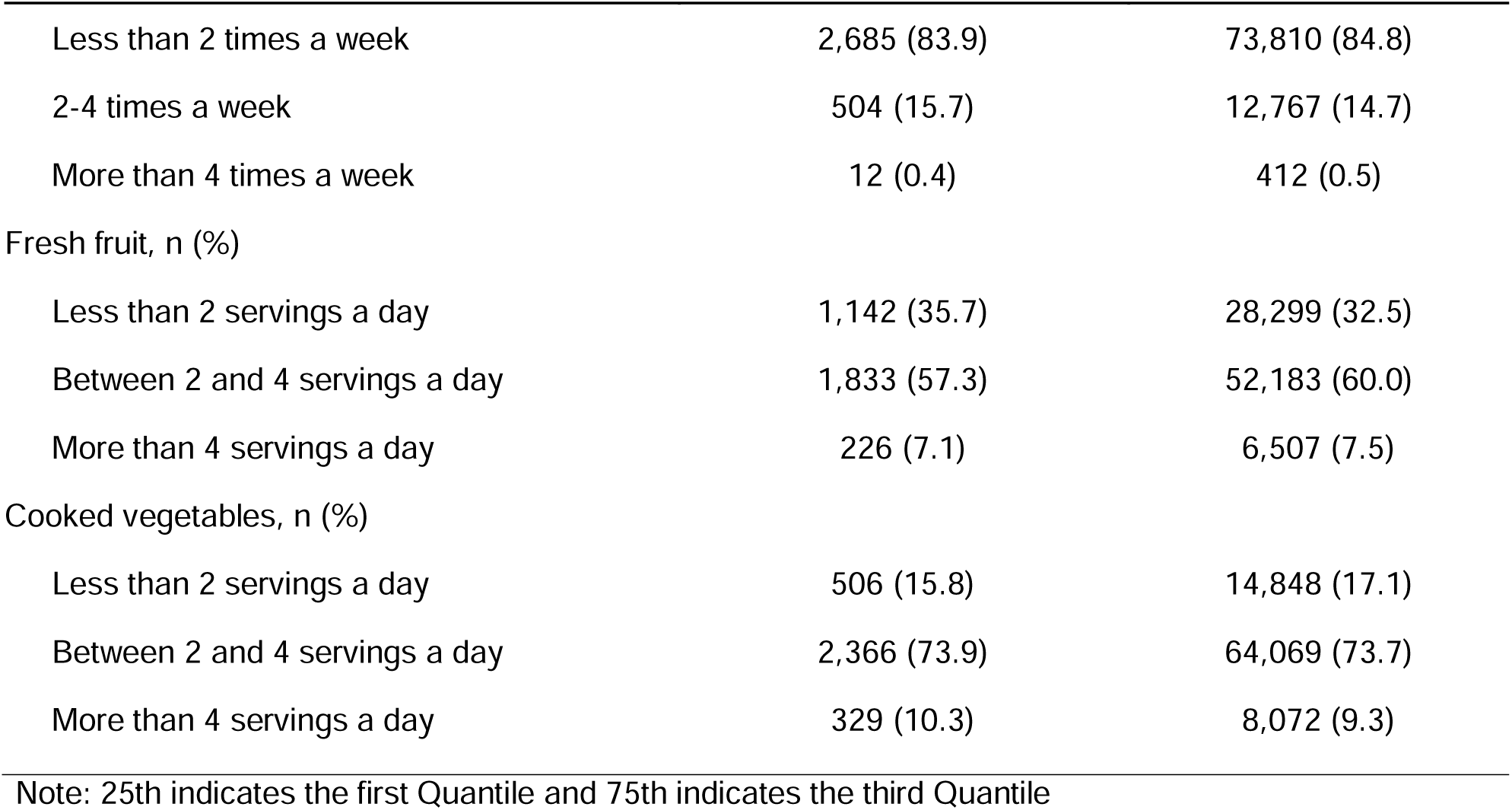
Distribution of participant characteristics by outcome status among adults in the UK Biobank accelerometer sub-cohort, 2013-2022.

| Baseline Characteristics | Cardiovascular disease<br>N = 3,298 | No cardiovascular<br>disease<br>N = 86,989 |
| --- | --- | --- |
| Follow up time in months, Median (25<sup>th</sup>, 75<sup>th</sup>) | 47.2 (17.2, 72.9) | 96.9 (90.3, 103.0) |
| Moderate-Vigorous Physical Activity:<br>Low-pass Filter Euclidean Norm Minus One,<br>Median (25<sup>th</sup>, 75<sup>th</sup>) | 375.8 (236.5, 535.5) | 466.9 (322.6, 644.4) |
| Moderate-Vigorous Physical Activity: Machine<br>learning, Median (25<sup>th</sup>, 75<sup>th</sup>) | 196.6 (81.5, 358.0) | 235.1 (116.0, 405.5) |
| Moderate-Vigorous Physical Activity: Activity<br>count, Median (25<sup>th</sup>, 75<sup>th</sup>) | 448.9 (268.1, 697.4) | 590.8 (376.7, 857.3) |
| Myocardial infarction, n (%) | 2,016 (63.0) | 0 (0) |
| Stroke, n (%) | 1,282 (40.0) | 0 (0) |
| Age, Median (25<sup>th</sup>, 75<sup>th</sup>) | 67.7 (62.7, 71.5) | 63.0 (55.9, 68.3) |
| Ethnicity, n (%) |  |  |
| Non-white | 83 (2.6) | 2,594 (3.0) |
| White | 3,118 (97.4) | 84,395 (97.0) |
| Sex, n (%) |  |  |
| Female | 1,200 (37.5) | 50,485 (58.0) |
| Male | 2,001 (62.5) | 36,504 (42.0) |
| Education, n (%) |  |  |
| None of the below | 422 (13.2) | 6,681 (7.7) |
| O levels/General Certificate of Secondary<br>Education or equivalent | 786 (24.6) | 21,315 (24.5) |
| A levels/Advanced Subsidiary, National<br>Vocational Qualifications/Higher National<br>Diploma/Higher National Certificate or<br>equivalent | 822 (25.7) | 20,486 (23.6) |
| College or University degree | 1,171 (36.6) | 38,507 (44.3) |
| Household income in British Pounds, n (%) |  |  |
| Less than 18,000 | 620 (19.4) | 10,948 (12.6) |
| 18,000 to 30,999 | 818 (25.6) | 18,831 (21.6) |
| 31,000 to 51,999 | 819 (25.6) | 22,832 (26.2) |
| 52,000 to 100,000 | 517 (16.2) | 20,374 (23.4) |
| Greater than 100,000 | 117 (3.7) | 5,955 (6.8) |
| Do not know/Prefer not to answer | 310 (9.7) | 8,049 (9.3) |
| Smoking, n (%) |  |  |
| Never | 1,529 (47.8) | 50,544 (58.1) |
| Previously | 1,368 (42.7) | 30,675 (35.3) |
| Currently | 304 (9.5) | 5,770 (6.6) |
| Alcohol consumption, n (%) |  |  |
| Never | 226 (7.1) | 4,728 (5.4) |
| Less than once a week | 637 (19.9) | 17,593 (20.2) |
| Once or twice a week | 765 (23.9) | 21,919 (25.2) |
| Three or four times a week | 793 (24.8) | 22,830 (26.2) |
| Daily or almost daily | 780 (24.4) | 19,919 (22.9) |
| Processed meat, n (%) |  |  |
| Less than 2 times a week | 2,136 (66.7) | 62,097 (71.4) |
| 2-4 times a week | 929 (29.0) | 21,832 (25.1) |
| More than 4 times a week | 136 (4.2) | 3,060 (3.5) |
| Red meat, n (%) |  |  |
| Less than 2 times a week | 435 (13.6) | 15,847 (18.2) |
| 2-4 times a week | 1,118 (34.9) | 29,458 (33.9) |
| More than 4 times a week | 1,648 (51.5) | 41,684 (47.9) |
| Oily fish, n (%) |  |  |
| Less than 2 times a week | 2,525 (78.9) | 71,962 (82.7) |
| 2-4 times a week | 646 (20.2) | 14,336 (16.5) |
| More than 4 times a week | 30 (0.9) | 691 (0.8) |
| Non-oily fish, n (%) |  |  |
| Less than 2 times a week | 2,685 (83.9) | 73,810 (84.8) |
| 2-4 times a week | 504 (15.7) | 12,767 (14.7) |
| More than 4 times a week | 12 (0.4) | 412 (0.5) |
| Fresh fruit, n (%) |  |  |
| Less than 2 servings a day | 1,142 (35.7) | 28,299 (32.5) |
| Between 2 and 4 servings a day | 1,833 (57.3) | 52,183 (60.0) |
| More than 4 servings a day | 226 (7.1) | 6,507 (7.5) |
| Cooked vegetables, n (%) |  |  |
| Less than 2 servings a day | 506 (15.8) | 14,848 (17.1) |
| Between 2 and 4 servings a day | 2,366 (73.9) | 64,069 (73.7) |
| More than 4 servings a day | 329 (10.3) | 8,072 (9.3) |

Note: 25th indicates the first Quantile and 75th indicates the third Quantile

We added several sensitivity analyses. First, we performed sex-stratified dose-response analysis to assess heterogeneity in protective associations. Second, we excluded cases that developed CVD outcomes within two years of accelerometer assessment to assess the presence of reverse causation. Third, we included type 2 diabetes mellitus, body mass index (BMI), self-reported depression, and self-reported health status as confounders, which we did not include in the main analysis, as they may mediate the association between PA and CVD. Fourth, we added clinical markers of CVD that may also serve as mediators rather than confounders. They are systolic and diastolic blood pressure, triglycerides, C-reactive protein, Hemoglobin A1c, high- and low-density lipoprotein, and the use of antiplatelet, antihypertensive, and lipid-lowering medications. Finally, we performed a competing risks analysis to account for the potential impact of non-CVD deaths.

The study was approved by the Faculty of Medicine Institutional Review Board (A08-M55-24A). We provided the Strengthening the Reporting of Observational studies in Epidemiology checklist (Supplementary File 2). Software codes to estimate dose-response associations using R statistical software and compute machine-learned MVPA from the validation data using the Accelerometer Python library are available (Supplementary File 3) (30).

## 3. Results

After excluding participants with insufficient wear time or low-quality accelerometer data (7,003 participants, 6.8% of the accelerometer sub-cohort), those with prior diagnoses of the outcome events before the study period (2,936; 2.9%), and those with missing behavioural risk factors (2,691; 2.6%), the final study sample comprised 90,237 participants (Supplementary eFigure 1). During follow-up, there were 2,016 and 1,282 incidences of first stroke and myocardial infarction before December 2022, respectively (Table 1). The median follow-up was 96.9 months (Interquartile Range [IQR]: 89.7-102.7) for CVD-free participants, and 47.2 (IQR: 17.2-72.9) for cases. Participants without CVD showed longer median MVPA durations, tended to be female and never smokers, had higher levels of education and income, and showed lower consumption of processed and red meat.

**Figure 1.**
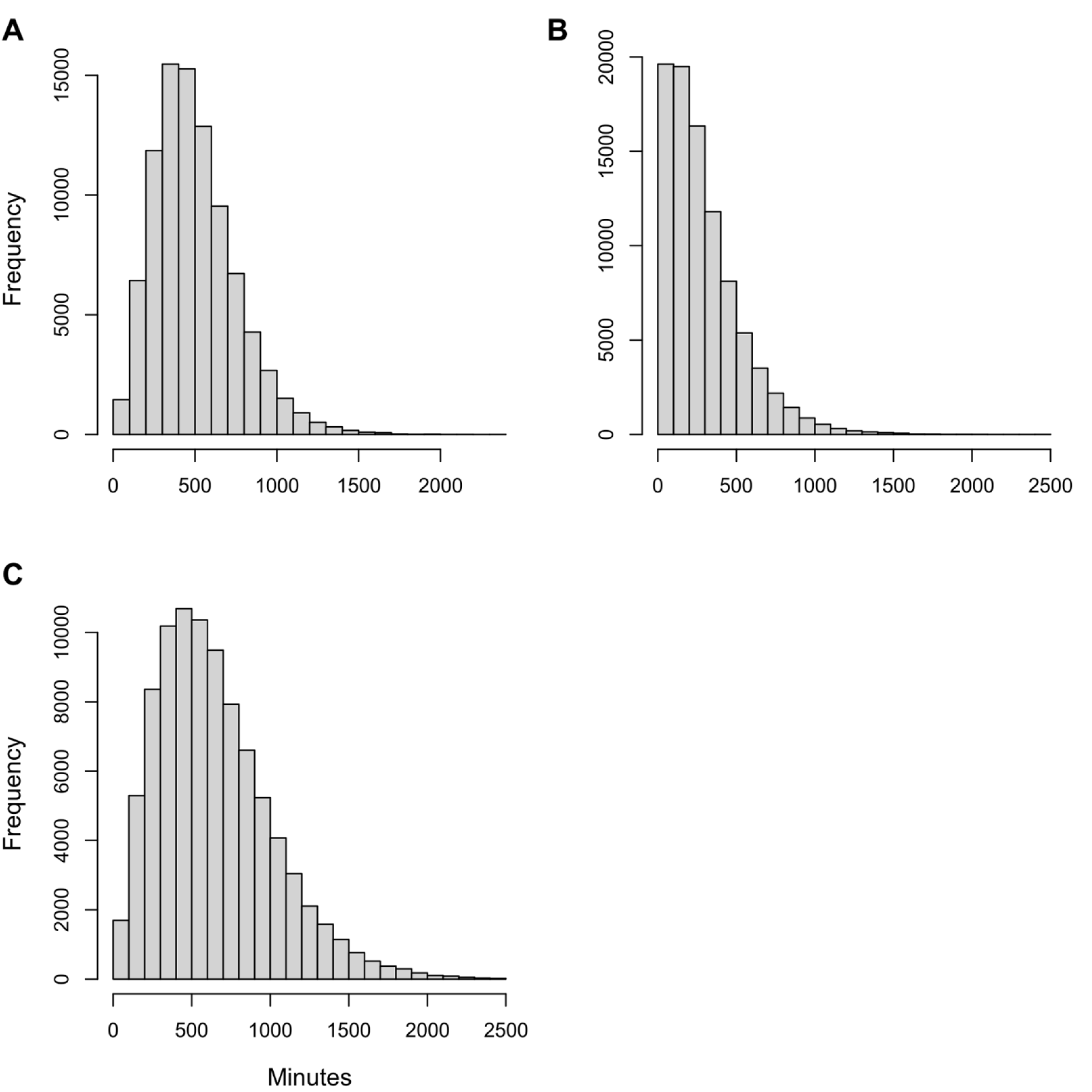
Distribution of accelerometer-derived weekly minutes spent for moderate-vigorous physical activity based on A) Low-pass Filter Euclidean Norm Minus One, B) machine learning, and C) activity count among adults in the UK Biobank accelerometer sub-cohort, 2013-2022.

Histograms of weekly MVPA duration showed varying skewness across the three metrics (Figure 1A-C). The median duration was the largest for the activity count-based metric (585.9 minutes/week, IQR: 372.1-853.2) and the smallest for machine-learning (234.1 minutes/week, IQR: 114.5-404.0). The agreement between the ground truth and the three MVPA metrics varied substantially, with machine learning showing the closest agreement (Figure 2A-C). The magnitude of disagreement appears to increase with duration. The corresponding Spearman’s coefficients are 0.50, 0.78 and 0.37 for LFENMO, machine-learning, and activity counts, respectively.

**Figure 2.**
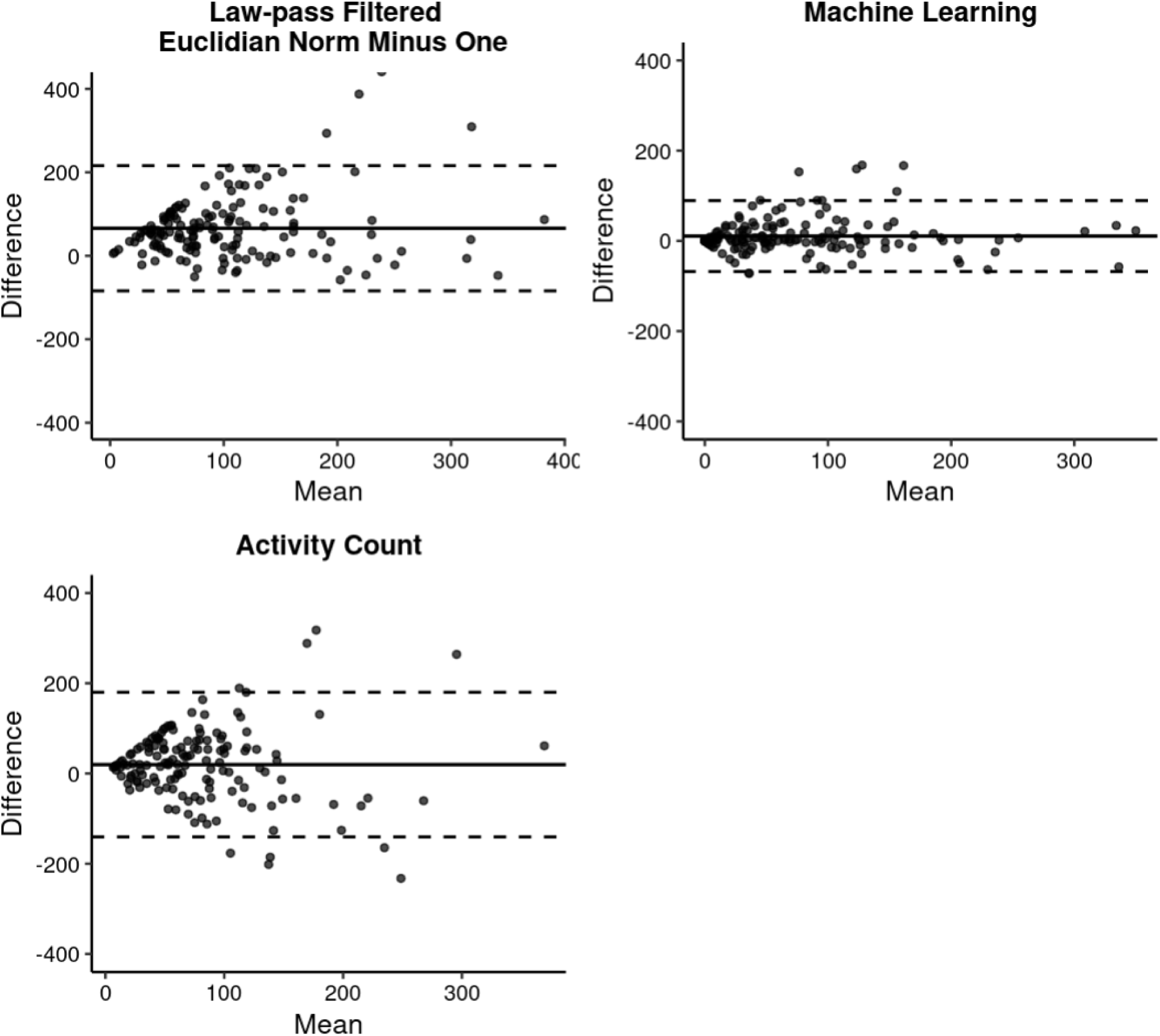
Bland-Altman plots showing agreement of the daily duration of moderate-vigorous physical activity between ground truth and three accelerometer-derived measures (X-axis) for Low-pass Filter Euclidian Norm Minus One (A), machine learning (B), and activity count (C), among the camera-recorded 24-hour activity validation study, consisting of 151 adult participants in Oxfordshire, United Kingdom, 2013-2022. Difference (y-axis) indicates pairwise difference, while Mean (x-axis) indicates pairwise mean.

The confounder-adjusted dose-response associations are shown in Figure 3A-F. The plots display associations up to 25 hours (1,500 minutes) per week, which is a high level of exposure with sparse data beyond this range. The unadjusted associations (Supplementary eFigure 2A-F) showed similar dose-response shapes to the adjusted associations, although the magnitude of the protective associations (reduction in Hazard Ratio [HR]) was more pronounced.

**Figure 3.**
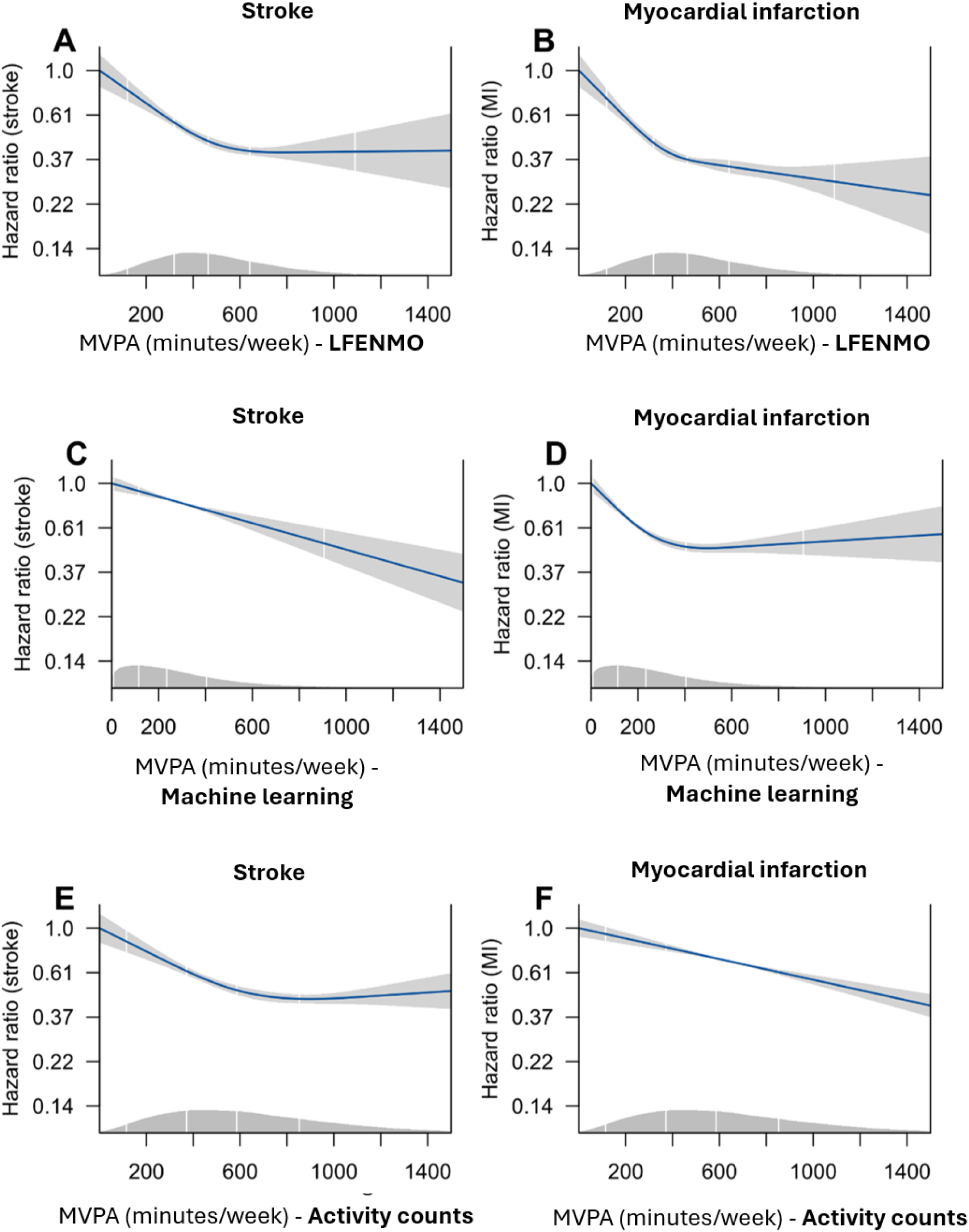
Dose-response associations between moderate-vigorous physical activity and stroke (left) and myocardial infarction (right) for Euclidean Norm Minus One (A and B), machine learning (C and D), and activity count (E and F) among adults in the UK Biobank accelerometer subcohort, 2013-2022. The histogram indicates the distribution of moderate-vigorous physical activity. Abbreviations: MVPA; Moderate-Vigorous Physical Activity, LFENMO; Low-pass Filter Euclidean Norm Minus One; MI, Myocardial Infarction.

For LFENMO, the association plateaued at approximately 650 minutes/week of MVPA of the inflection point for both outcomes (Figures 3A and 3B for stroke and myocardial infarction, respectively). For machine-learning, the protective association was linear: the model without spline showed a better model fit according to Akaike’s Information Criterion (Figure 3C). For myocardial infarction, machine-learning showed a curvilinear association with a very modestly increased HR after the inflection point at approximately 400 minutes/week of MVPA, i.e., *J-shape* (Figure 3D). However, this increased trend is inconclusive given the width of the 95% Confidence Interval (CI). For activity counts, the estimated curvilinear association with stroke is similar to that of LFENMO, but the inflection point was slightly larger at approximately 800 minutes/week (Figure 3E). For myocardial infarction, activity counts showed a linear association, unlike LFENMO and machine-learning (Figure 3F). Regarding the magnitude of the protective effect, LFENMO showed the strongest reduction of HR down to approximately HR=0.37 in both outcomes.

For the sensitivity analysis focusing on sex-specific analysis of stroke, the shape of association was similar between females (Supplementary eFigures3-5A, green shade) and males (Supplementary eFigures 3-5C, grey shade) for all three accelerometer-derived metrics (Supplementary eFigures 3, 4, and 5 for LFENMO, machine-learning, and activity count, respectively). However, the protective association was slightly stronger (with slightly lower HR values) for males. For myocardial infarction, females had distinct shapes of associations with a more pronounced and linear decrease in HR, compared to males with more curvilinear associations, for all three metrics (contrast of supplementary eFigures 3-5B for females vs. eFigures 3-5D for males). The exclusion of participants who developed the outcomes within 2 years of the exposure assessment did not yield noticeable changes. Also, the associations remained robust after accounting for potential mediators (BMI, depression, and type 2 diabetes mellitus) and clinical markers of CVD, as well as medications. Finally, competing risk analysis showed a more pronounced protective effect of MVPA for both stroke and myocardial infarction (Supplementary eFigure 6). However, the overall shape of the associations for all MVPA metrics remained very similar.

## 4. Discussion

We investigated the comparability of the dose-response association between MVPA and CVD across three MVPA metrics from accelerometer data using the UK Biobank accelerometer sub-cohort. Unlike LFENMO and activity count metrics, machine-learned MVPA showed a curvilinear association with stroke. On the other hand, only the activity count metric showed a linear protective association of MVPA with myocardial infarction.

Agreement analysis indicates that machine learning had the highest measurement error relative to the ground truth, followed by FEENMO, while activity counts showed the least agreement. Previous studies have found higher accuracy for machine learning in classifying physical activities than for cut-point methods, such as ENMO and activity counts (15,19,22,31). In our data, all three accelerometer-MVPA metrics led most participants to achieve more than 150 minutes/week of MVPA, with LFENMO and activity counts showing higher durations. However, the amount of MVPA reported by cut-point methods depends entirely on the chosen threshold (i.e., cut-point) for MVPA (14).

The strongest agreement of machine-learning with the ground truth is expected, since the model we used in this study was trained on the same validation (Capture-24) data, (22), whereas cut-point methods, including ours, utilize threshold values for MVPA calibrated and validated by external data (15,16,18). Unsurprisingly, machine-learning previously showed a reduced performance when externally generalized to another study population, even for laboratory-generated (non-free-living) data (32). However, the recent availability of foundation models and methods to fine-tune these models with a minimum amount of cohort-specific internal validation data may represent the advantage of machine-learning over methods based on cut-point (33).

The three accelerometer metrics agreed that increasing MVPA incrementally reduces the risk of both outcomes, even when the duration was below the recommended 150 minutes/week cutoff, consistent with previous meta-analyses using self-report (8–10). However, the shape of the associations (steepness and plateau point) varies noticeably across metrics. Specifically, only the machine-learned MVPA showed a linear decrease of HR for stroke (as opposed to the curvilinear associations for LFENMO and activity counts). For myocardial infarction, activity counts showed a linear protective association, while the other two showed a curvilinear.

A previous meta-analysis examining accelerometer-derived physical activity metrics and all-cause mortality reported a curvilinear dose-response association (13). While dose-response meta-analyses for CVD outcomes specific to accelerometer-derived physical activity are lacking, those based on self-reported measures suggest a curvilinear association for both stroke and myocardial infarction (8,9). On the other hand, individual studies focusing on accelerometer-derived physical activity have reported both linear and curvilinear associations (11,12). Our study indicates that methodological choices in accelerometer processing may partly explain variations in dose-response associations, calling for careful interpretation and comparison of associations estimated in studies that use a single processing method. For this reason, as previously suggested, researchers are encouraged to report associations using multiple accelerometer processing methods for informed comparison, where possible (31). We note that the superior agreement of machine learning with the ground truth does not necessarily imply that the associations based on this method are most credible, as the machine-learning model was trained on the ground-truth data (not externally generalized, as in cut-point methods).

Our sensitivity analysis indicates that for the female subsample, the greater protective effect of MVPA for myocardial infarction, as agreed by all three metrics, is consistent with a previous study (9). However, we did not observe any female-specific benefit in stroke outcome, again agreed by all metrics.

The strengths of this study include comparing three accelerometer processing metrics to address the unexplored discrepancy in dose-response associations. This is a novel addition to the literature, which has focused on comparing the prevalence of physical activity across accelerometer processing methods among children, rather than on dose-response associations among adults (20,34). The limitations include the UK Biobank’s lack of representativeness of the UK population, since the sampling is participant-driven. Nevertheless, a previously reported association between meeting the physical activity guideline and CVD incidence did not differ statistically before and after post-stratification, suggesting the potential absence of selection bias (35). Also, we did not assess variation of associations due to other influential factors, such as wearable device types (e.g., Axivity v.s. ActiGraph), size of epoch, intensity thresholds (e.g., use of 125mg cut-point instead of 100mg for LFENMO), type of vector magnitude summary (e.g., Monitor-Independent Movement Summary, instead of LFENMO), and location of device e.g., hip vs. wrist, all of which contribute to the variation of findings (14,15,31).

Thus, future work should include a more comprehensive comparison of associations across these methodological factors. Such comparison studies should ideally include various machine-learning algorithms trained by data from free-living participants, with external generalization to other cohorts for fair comparison with cut-point methods. Another research avenue is to investigate dose-response associations for other PA intensities, including light physical activity, total physical activity (combination of light and MVPA), and sedentary behaviour. Finally, studies to calibrate dose-response associations using measurement error models and non-linear survival analysis, incorporating ground-truth (internal validation) data, should be conducted (8). This is because even the machine-learning model showed substantial measurement error in our study, which should be accounted for.

## 5. Conclusions

We demonstrated that the selection of accelerometer processing methods for MVPA can influence the dose-response association between MVPA and CVD. Thus, when characterizing the benefits of MVPA for CVD prevention, researchers should exercise caution in interpreting findings from a single MVPA measure.

## Data availability statement

UK Biobank data are available for research upon approval from the UK Biobank Access Management System.

## Description of conflicts of interest

Sharma has received research support from Roche Diagnostics, Boehringer Ingelheim, Novartis, Janssen, Novo Nordisk, Servier, AstraZeneca, and Takeda. Also, Sharma has equity stake and/or has co-founded: AREA19, Aerocardia, Perc-assist.

## Declaration of generative AI in scientific writing

We did not use generative AI in any stage of manuscript preparation.

## Authorship contributions

**Lapointe**: Conceptualization, Data curation, Formal Analysis, Visualization, Methodology, Investigation, Writing – original draft, Writing – review & editing. **Kapur**: Conceptualization, Software, Data Curation, Investigation, Writing – review & editing. **Panter**: Conceptualization, Methodology, Investigation, Writing – review & editing. **Fuller**: Conceptualization, Methodology, Investigation, Writing – review & editing. **Sharma**: Conceptualization, Methodology, Interpretation, Validation, Writing – review and editing. **Mamiya**: Conceptualization, Supervision, Funding acquisition, Project administration, Methodology, Interpretation, Writing – review & editing.

## Supporting information

STROBE Checklist

## Data Availability

All data are available for research upon approval from the UK Biobank Access Management System.

## Acknowledgements

We appreciate Dr. Shing Chan and Dr. Aiden Doherty for their helpful comments on the Capture-24 dataset and the Accelerometer Python library.

