## Supplementary Figures for "Investigating the comparability of wearable accelerometer methods in the association between physical activity and cardiovascular disease: a cohort study using UK Biobank"

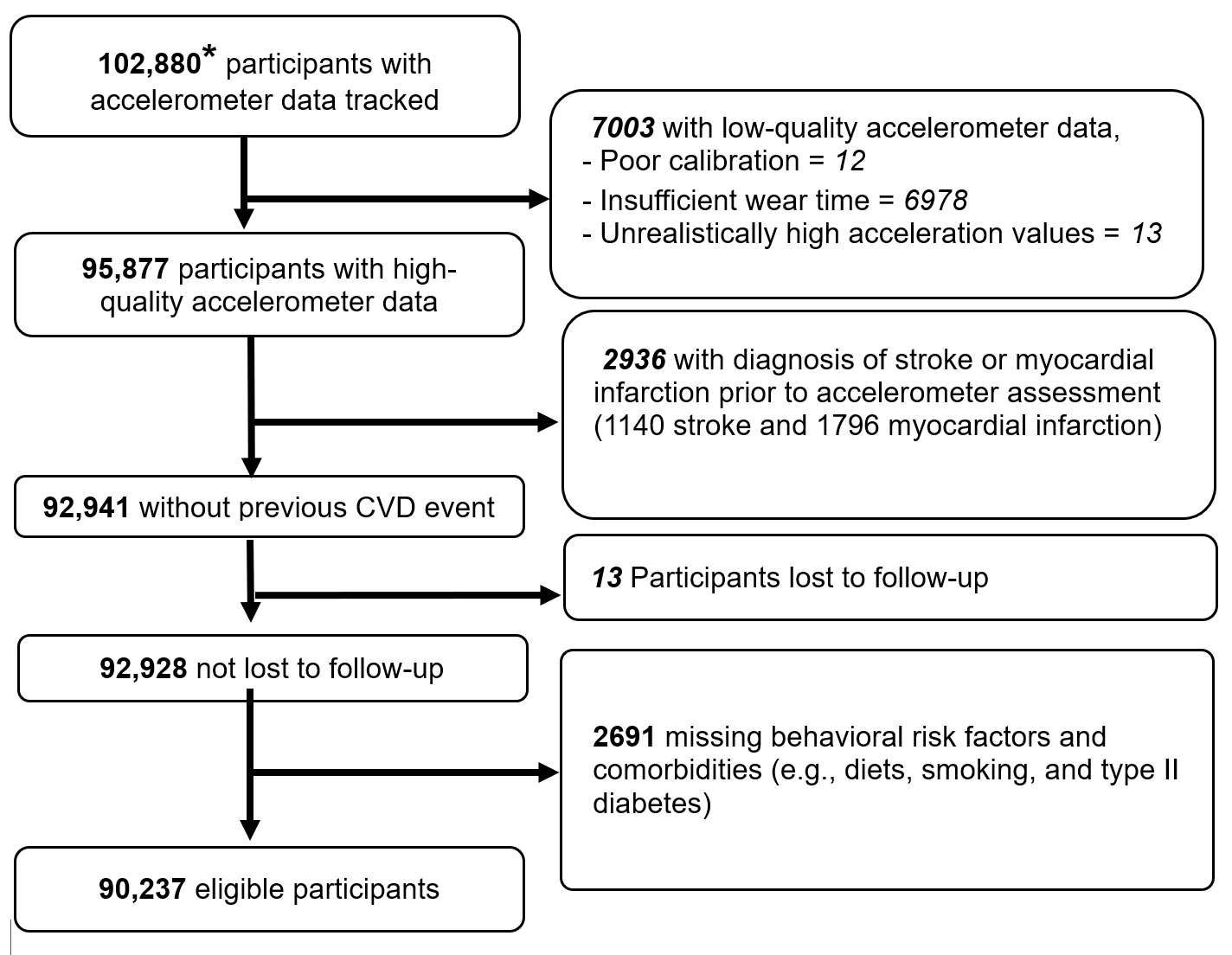
Supplementary eFigure 1. Flowchart describing the exclusion process for adults in the UK Biobank accelerometer sub-cohort, 2013-2022.

*This is the remaining number after the UK Biobank removed approximately 1200 individuals who requested to remove their entire data from the cohort.

Abbreviation: CVD, Cardiovascular disease

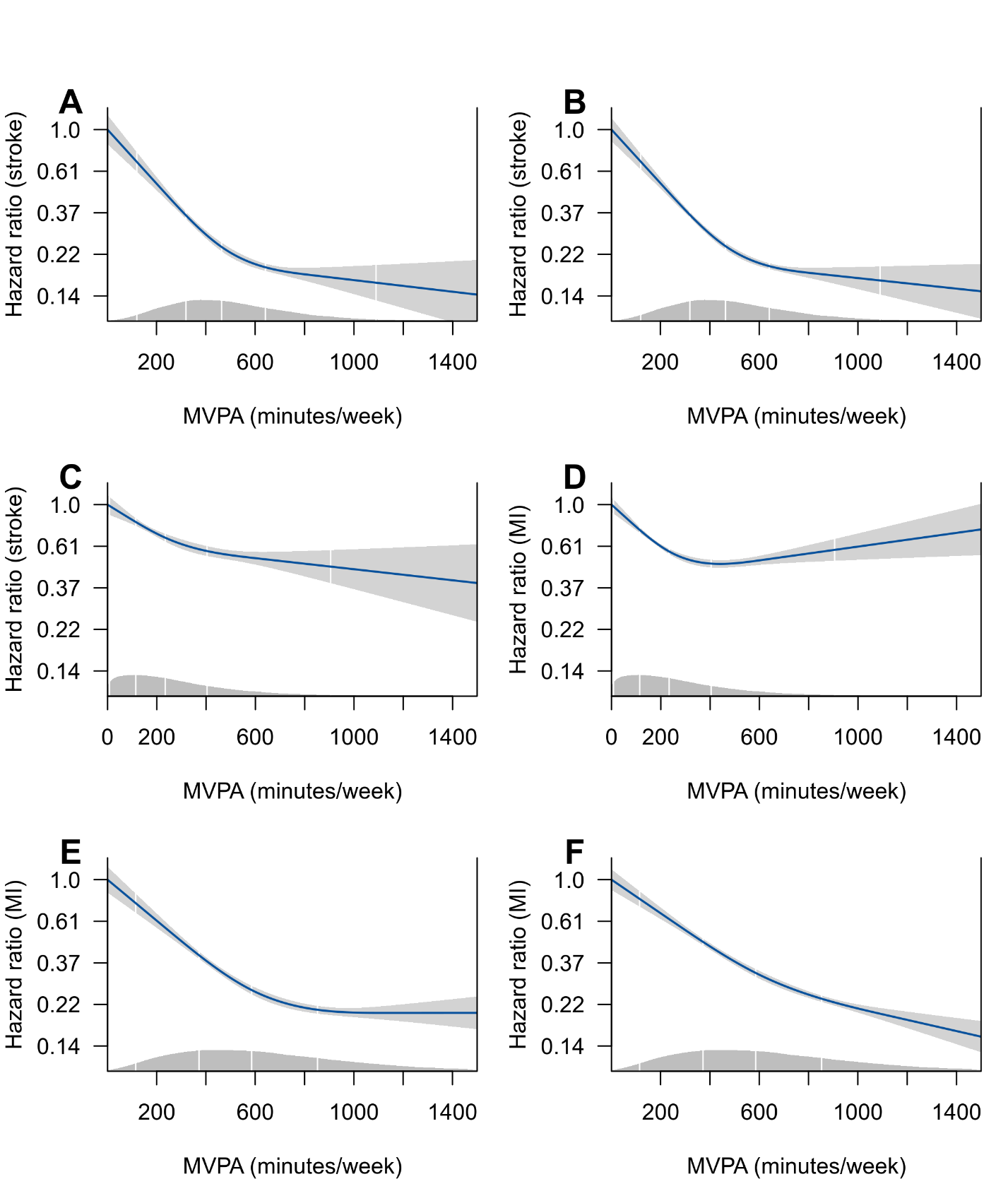

Supplementary eFigure 2. Unadjusted dose-response associations between moderate-vigorous physical activity and stroke (left) and myocardial infarction (right) for Low-pass Filter Euclidean Norm Minus One (A and B), machine learning (C and D), and activity count (E and F) among adults in the UK Biobank accelerometer sub-cohort, 2013-2022. The histogram indicates the distribution of moderate-vigorous physical activity.

Abbreviations: MVPA; Moderate-Vigorous Physical Activity, MI; Myocardial infarction

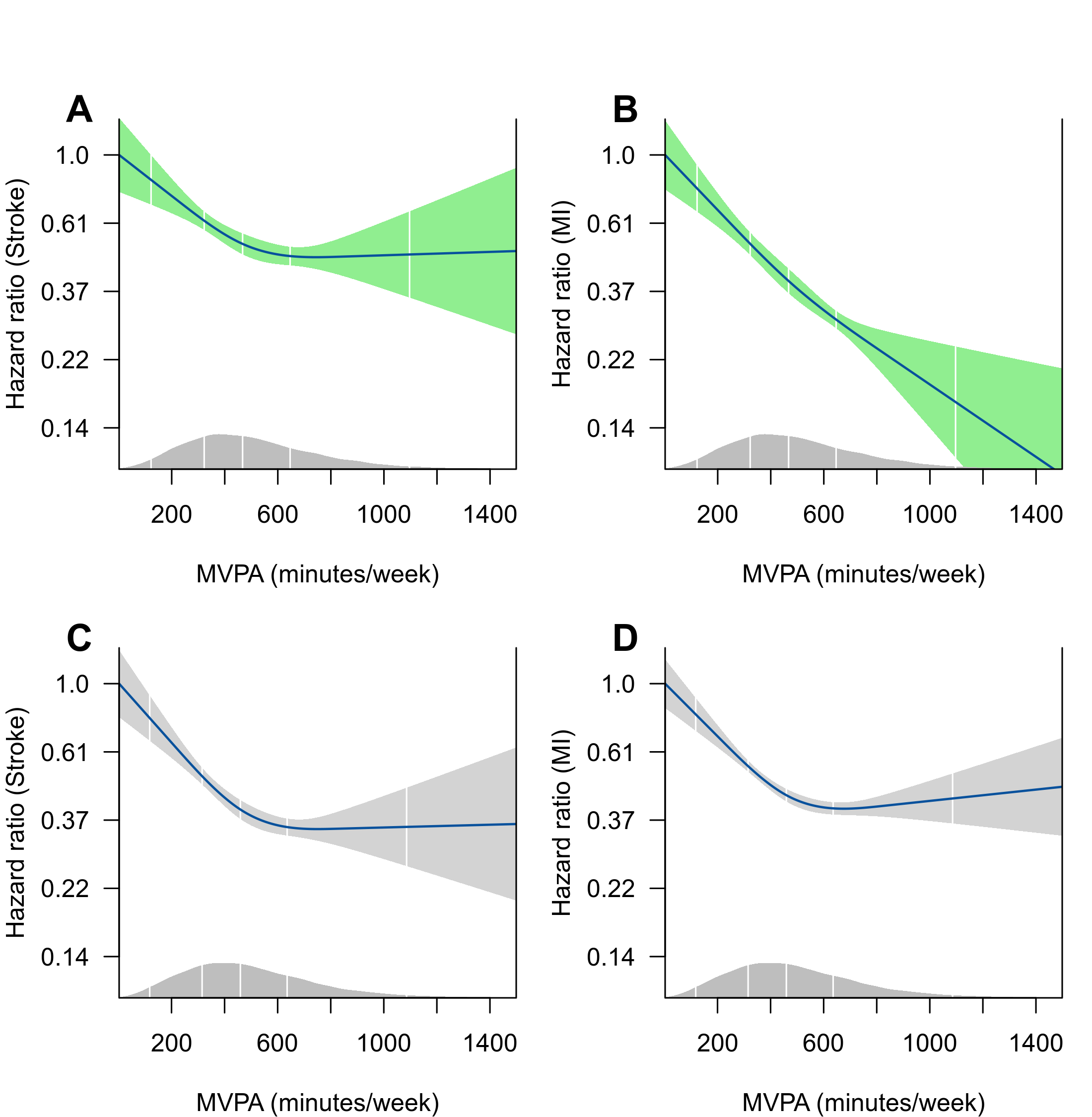

### Supplementary eFigure 3. Dose-response association between Moderate-Vigorous Physical Activity computed by Euclidean Normal Minus One and stroke (A) and myocardial infarction (B) among females, for stroke (C) and myocardial infarction (D) among males. Data are from adults in the UK Biobank accelerometer sub-cohort, 2013-2022. The green band indicates estimation for females.

Abbreviations: MVPA; Moderate-Vigorous Physical Activity, ENMO; Myocardial Infarction, MI

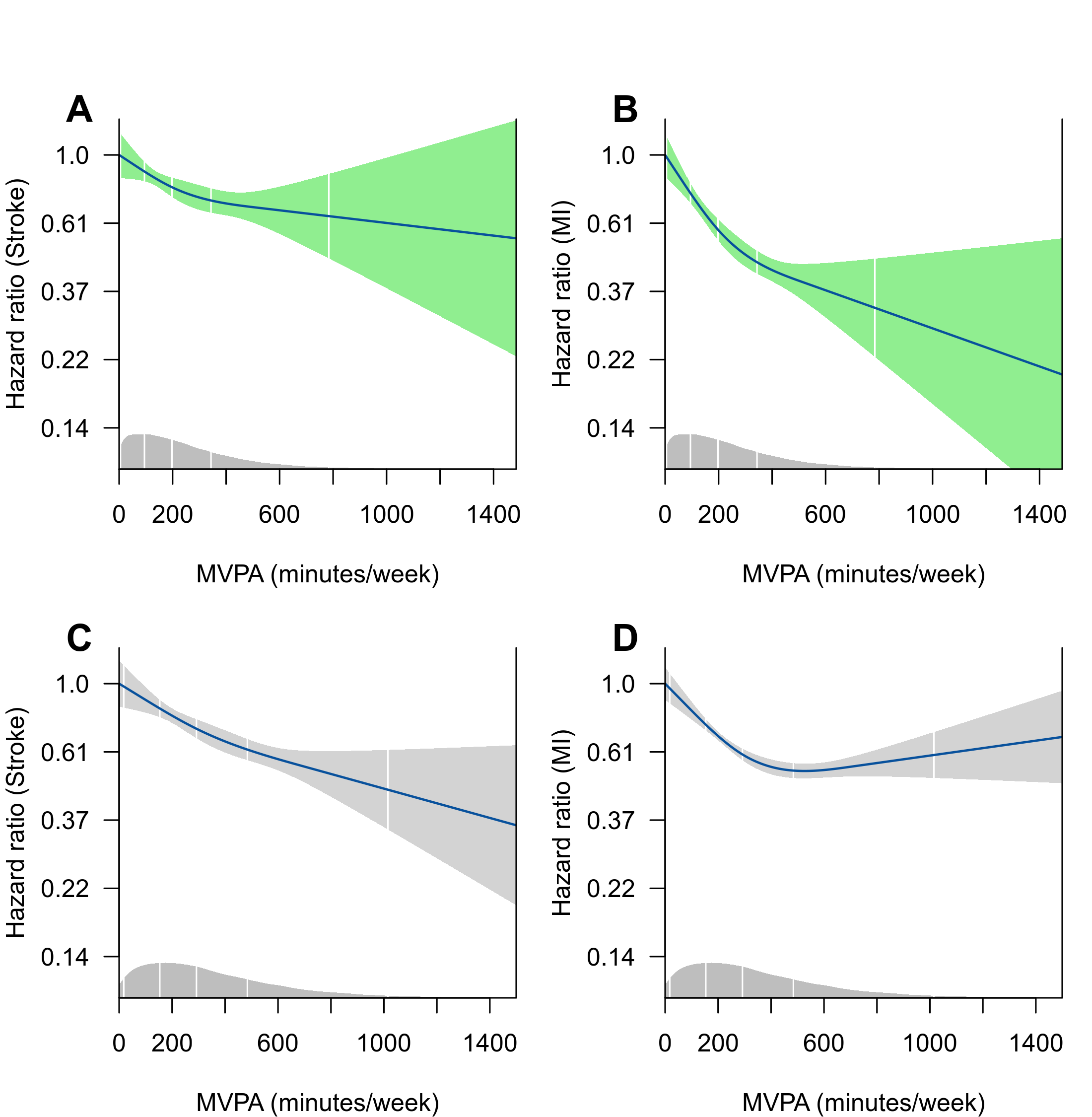

### Supplementary eFigure 4. Dose-response association between Moderate-Vigorous Physical Activity estimated by machine learning and stroke (A) and myocardial infarction (B) among females, for stroke (C) and myocardial infarction (D) among males. Data are from adults in the UK Biobank accelerometer sub-cohort, 2013-2022. The green band indicates the estimated value for females.

Abbreviations: MVPA; Moderate-Vigorous Physical Activity, ENMO; Myocardial Infarction, MI

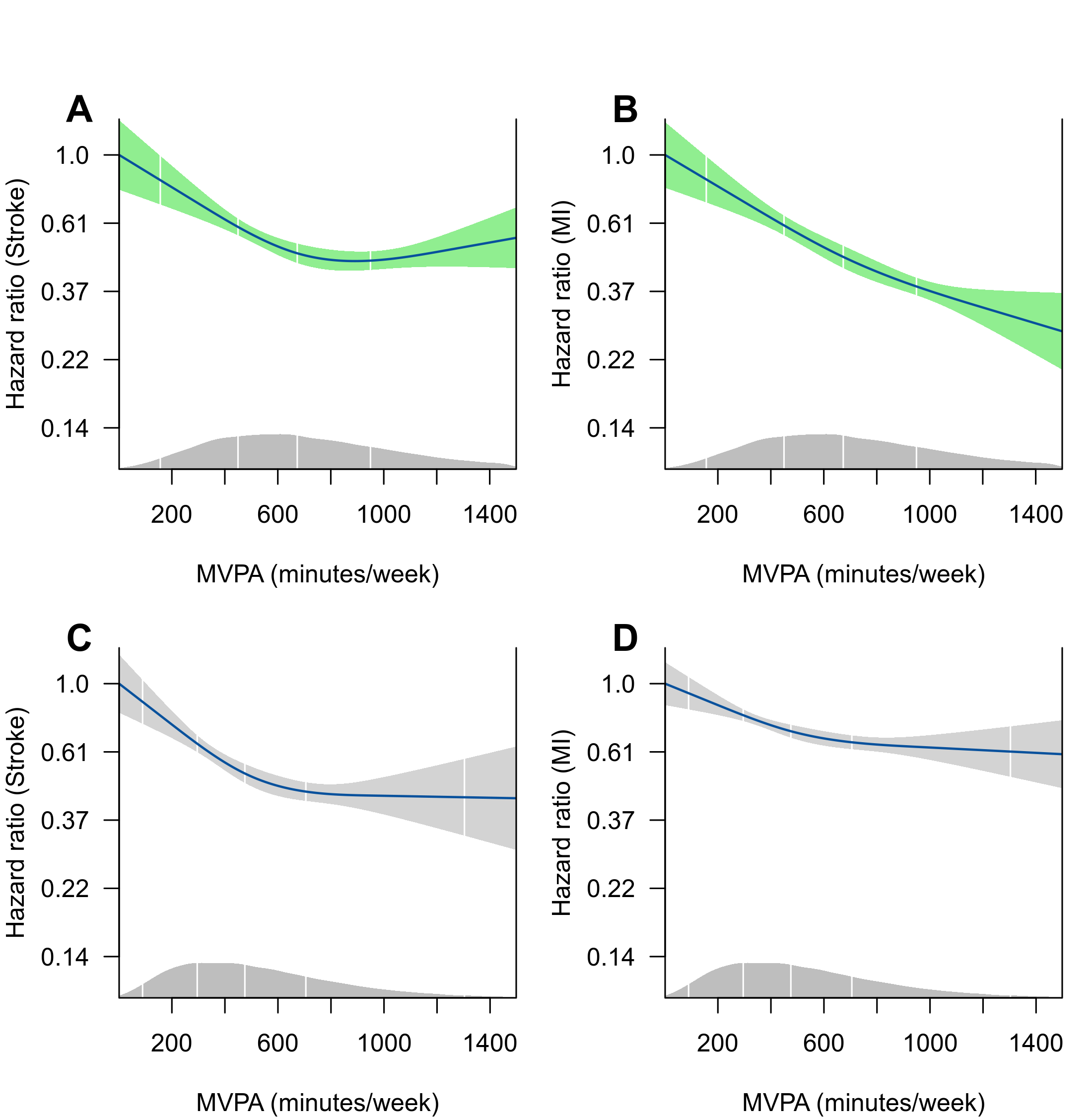

### Supplementary eFigure 5. Dose-response association between Moderate-Vigorous Physical Activity computed by activity count and stroke (A) and myocardial infarction (B) among females and stroke (C) and myocardial infarction (D) among males. Data are from adults in the UK Biobank accelerometer sub-cohort, 2013-2022. The green band indicates estimation for females.

Abbreviations: MVPA; Moderate-Vigorous Physical Activity, MI; Myocardial Infarction

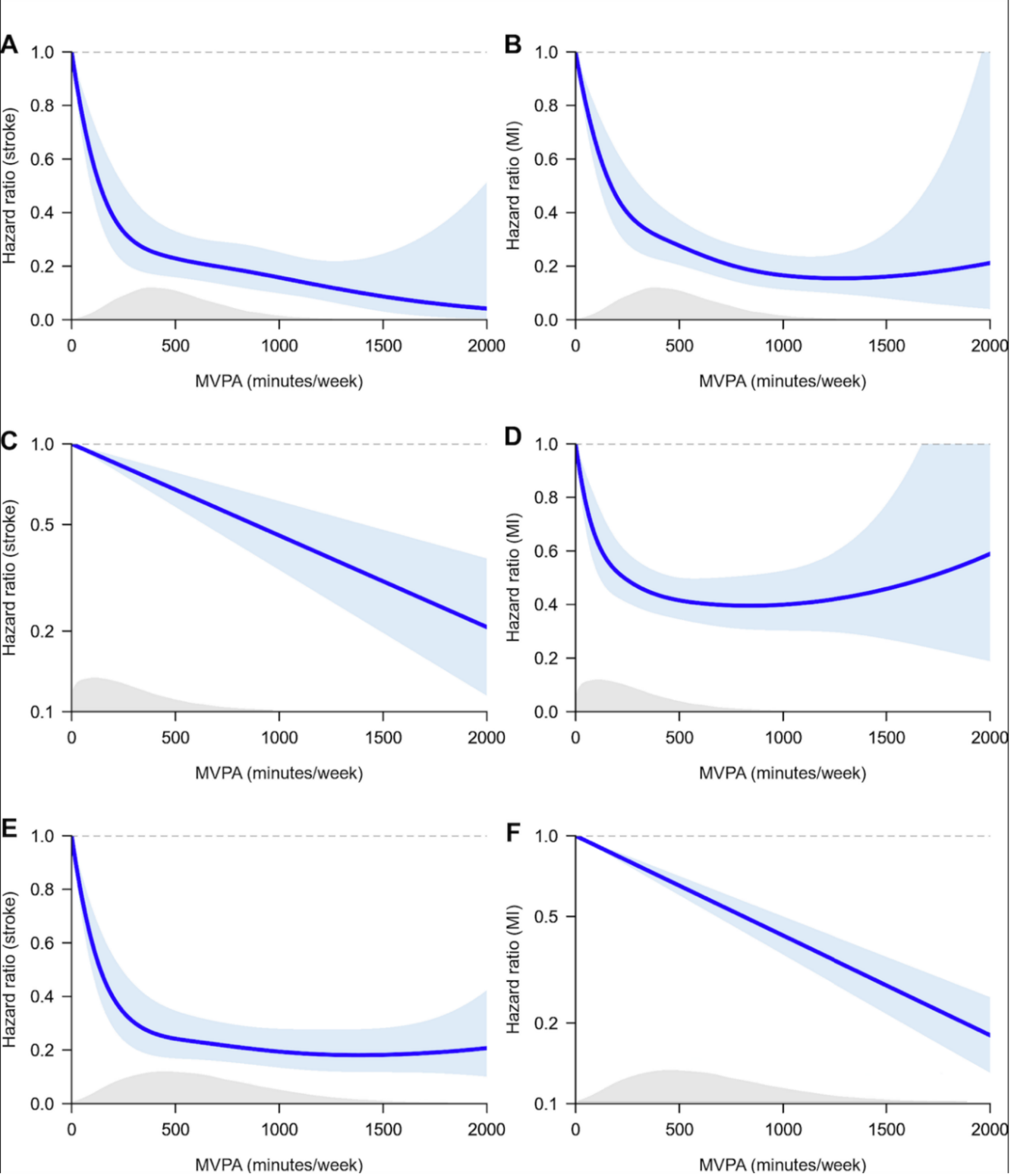

### Supplementary eFigure 6 - Dose-response associations after accounting for competing risk due ot non-cardiovascular deaths, for stroke (left) and myocardial infarction (right), for Euclidian Normal Minus One (A and B), machine learning (C and D), and activity count (E and F) among adults in the UK Biobank accelerometer sub-cohort, 2013-2022.

### Note that the x-axis (MVPA duration) is extended to 2000 minutes. For plots C and F, model selection based on Akaike’s information criterion indicated a superior model with the linear exposure-outcome associations, rather than a non-linear (spline-based) model.

Abbreviations: MVPA; Moderate-Vigorous Physical Activity; Myocardial Infarction, MI
